## Supplementary figures and images for "Separating Newborns from Mothers and Maternal Consent for Newborn Care and the Association with Health Care Satisfaction, Use and Breastfeeding: Findings from a longitudinal survey in Kenya"

### Supplement 1

#### Supplement 1. Flowchart of baseline and follow-up analytic sample sizes

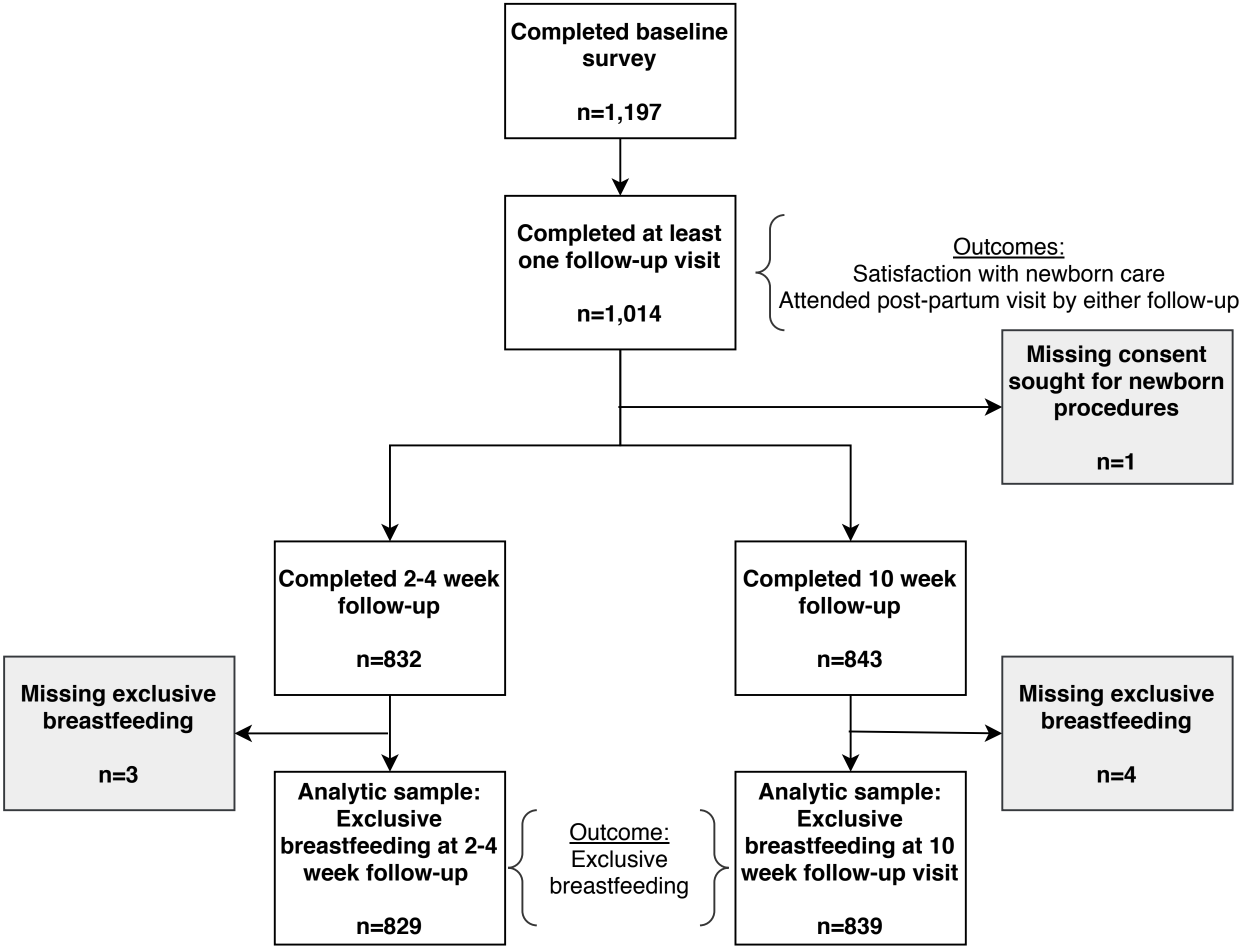
