## Supplement 2 for "Separating Newborns from Mothers and Maternal Consent for Newborn Care and the Association with Health Care Satisfaction, Use and Breastfeeding: Findings from a longitudinal survey in Kenya"

**Supplement 2. Distributions of clinical and breastfeeding care indicators by reports of newborn separation and newborn consent indicators, (n=1,014)**

| Variable | Total | Newborn separation: Not separated | Newborn separation: Separated | Newborn separation: p-value | Newborn separation >10min: Not separated or separated ≤ 10min | Newborn separation >10min: Separated >10min | Newborn separation >10min: p-value | Consent sought for newborn procedures[1]: Did not ask for permission | Consent sought for newborn procedures[1]: Asked for permission | Consent sought for newborn procedures[1]: p-value |
|---|---|---|---|---|---|---|---|---|---|---|
| Total number in group | 1,014 | 836 | 178 | | 886 | 128 | | 607 | 406 | |
| **Clinical quality of care indicators** | | | | | | | | | | |
| Was your baby put on your abdomen or chest as soon as it was born? | | | | | | | | | | |
| No | 13.6% | 13.0% | 16.3% | 0.467 | 13.0% | 18.0% | 0.287 | 15.8% | 10.3% | 0.032 |
| Yes | 86.3% | 86.8% | 83.7% | | 86.9% | 82.0% | | 84.0% | 89.7% | |
| Don't know | 0.1% | 0.1% | 0.0% | | 0.1% | 0.0% | | 0.2% | 0.0% | |
| Within two hours of delivery, did a health provider examine your baby? | | | | | | | | | | |
| No | 25.0% | 25.7% | 21.9% | 0.049 | 26.4% | 15.6% | 0.001 | 31.3% | 15.5% | <0.001 |
| Yes | 74.1% | 73.7% | 75.8% | | 73.0% | 81.2% | | 67.5% | 84.% | |
| Don't know | 0.8% | 0.6% | 1.7% | | 0.6% | 2.3% | | 1.2% | 0.2% | |
| Missing | 0.1% | 0.0% | 0.6% | | 0.0% | 0.8% | | 0.0% | 0.2% | |
| Was your baby wiped dry within a few minutes after birth? | | | | | | | | | | |
| No | 7.4% | 7.3% | 7.9% | 0.013 | 7.2% | 8.6% | 0.091 | 9.2% | 4.7% | 0.003 |
| Yes | 90.4% | 91.1% | 87.1% | | 91.0% | 86.7% | | 88.0% | 94.3% | |
| Don't know | 2.2% | 1.6% | 5.1% | | 1.8% | 4.7% | | 2.8% | 1.0% | |
| Has your baby been bathed yet? | | | | | | | | | | |
| No | 92.6% | 93.3% | 89.3% | <0.00 | 93.3% | 87.5% | <0.00 | 92.6% | 92.6% | 0.723 |
| Yes | 5.8% | 5.9% | 5.6% | | 5.9% | 5.5% | | 5.6% | 6.2% | |
| Don't know | 1.6% | 0.8% | 5.1% | | 0.8% | 7.0% | | 1.8% | 1.2% | |
| After birth, did any health care provider examine the cord? | | | | | | | | | | |
| No | 28.0% | 27.4% | 30.9% | 0.344 | 27.9% | 28.9% | 0.809 | 34.8% | 18.0% | <0.001 |
| Yes | 72.0% | 72.6% | 69.1% | | 72.1% | 71.1% | | 65.2% | 82.0% | |
| After birth, did any health care provider assess the temperature of your baby? | | | | | | | | | | |
| No | 51.1% | 52.6% | 43.8% | 0.033 | 52.0% | 44.5% | 0.113 | 56.3% | 43.1% | <0.001 |
| Yes | 48.9% | 47.4% | 56.2% | | 48.0% | 55.5% | | 43.7% | 56.9% | |
| After birth, did any health care provider counsel you on danger signs for newborns? | | | | | | | | | | |
| No | 63.4% | 63.4% | 63.5% | 0.983 | 63.4% | 63.3% | 0.974 | 73.0% | 49.0% | <0.001 |
| Yes | 36.6% | 36.6% | 36.5% | | 36.6% | 36.7% | | 27.0% | 51.0% | |
| **Breastfeeding care indicators** | | | | | | | | | | |
| Within two hours of delivery, did a health provider check if breastfeeding was going well. | | | | | | | | | | |
| No | 24.2% | 22.8% | 30.3% | 0.048 | 23.9% | 25.8% | 0.248 | 29.8% | 15.8% | <0.001 |
| Yes | 75.6% | 77.0% | 69.1% | | 76.0% | 73.4% | | 69.9% | 84.2% | |
| Don't know | 0.2% | 0.1% | 0.6% | | 0.1% | 0.8% | | 0.3% | 0.0% | |
| While you were in the hospital for the delivery of this baby did any one counsel/talk to you about breastfeeding? | | | | | | | | | | |
| No | 34.6% | 33.5% | 39.9% | 0.103 | 34.% | 39.1% | 0.258 | 40.7% | 25.4% | <0.001 |
| Yes | 65.4% | 66.5% | 60.1% | | 66.% | 60.9% | | 59.3% | 74.6% | |
| While you were in the hospital for the delivery of this baby did any one help you with breastfeeding by observing or showing you how to breastfeed? | | | | | | | | | | |
| No | 42.4% | 40.6% | 50.9% | 0.013 | 41.5% | 48.8% | 0.126 | 51.1% | 29.6% | <0.001 |
| Yes | 57.6% | 59.4% | 49.1% | | 58.5% | 51.2% | | 48.9% | 70.4% | |

Note: For analyses, clinical quality and breastfeeding care indicators, “don’t know” responses were recoded as “no,” and one missing response was conservatively recoded as “yes.”
